# Use of glycolysis enhancing drugs has less risk of Parkinson’s disease than 5α-reductase inhibitors

**DOI:** 10.1101/2022.04.01.22273312

**Authors:** Jacob E. Simmering, Michael J. Welsh, Jordan Schultz, Nandakumar S. Narayanan

## Abstract

**Background:** Terazosin and closely related α1-adrenergic receptor antagonists (doxazosin and alfuzosin; TZ/DZ/AZ) enhance glycolysis and reduce neurodeneration in animal models. Observational evidence in humans from several databases support this finding; however, a recent study has suggested that tamsulosin, the comparator medication, increases risk of Parkinson’s disease. We consider a different comparison group of men taking 5α-reductase inhibitors as a new, independent comparison allowing us to both obtain new estimates of the association between TZ/DZ/AZ and Parkinson’s disease outcomes and validate tamsulosin as an active comparator.

**Methods:** Using the Truven Health Analytics Marketscan database, we identified men without Parkinson’s disease, newly started on TZ/DZ/AZ, tamsulosin, or 5α-reductase inhibitors. We followed these matched cohorts to compare the hazard of developing Parkinson’s disease.

**Results:** We found that men taking TZ/DZ/AZ had a lower hazard of Parkinson’s disease than men taking tamsulosin (HR=0.72, 95% CI: 0.66-0.78, n=239,946) and lower than men taking a 5α-reductase inhibitors (HR=0.81, 95% CI: 0.72-0.92, n=129,320). The hazard for men taking tamsulosin was not statistically significantly different than for men taking 5α-reductase inhibitors (HR=1.10, 95% CI: 1.00-1.22, n=157,490).

**Conclusions:** These data suggest that men using TZ/DZ/AZ have a lower risk of developing Parkinson’s disease than those using tamsulosin or 5α-reductase inhibitors while users of tamsulosin and 5α-reductase inhibitors have relatively similar survival functions.

## Introduction

Parkinson’s disease (PD) is a neurodegenerative disease with increasing overall prevalence as the population ages. (1, 2) Although the causes of sporadic PD remain uncertain, there is substantial evidence implicating altered energy metabolism. Diverse genetic causes of PD including *PINK1, parkin, DJ-1, CHCHD2, alpha-synuclein*, and *LRRK2* mutations (3, 4) impair energy metabolism, and the mitochondrial toxins MPTP and rotenone induce PD. (2) Aging, the primary risk factor for PD, is associated with reduced brain energy metabolism, lower rates of mitochondrial biogenesis, and lower brain adenosine triphosphate (ATP) levels. (2, 5-7) These diverse observations converge on impaired brain energy metabolism as a common pathogenic mechanism of PD. Consequently, enhancing energy metabolism might slow or prevent PD.

Indeed, recent data suggest that enhancing glycolysis slows neurodegeneration. Specifically, terazosin and the related α1-adrenergic receptor antagonists doxazosin and alfuzosin (collectively TZ/DZ/AZ) bind to and activate phosphoglycerate kinase 1 (PGK1), the first ATP-producing enzyme in glycolysis. In genetic and toxin-induced models of PD in mice, rats, flies and induced pluripotent stem cells from PD patients, TZ/DZ/AZ increased brain and cellular ATP levels and prevented or slowed neuron loss. (8) TZ also increased brain ATP in a small pilot study in humans. (9) In clinical practice, TZ/DZ/AZ are routinely used to treat benign prostatic hyperplasia (BPH), a common condition in older men. Tamsulosin, another α1-adrenergic receptor antagonist, has similar clinical indications and effectiveness (10) but does not engage PGK1. (8) This creates natural comparison groups of men with similar health status who are electing to undergo pharmaceutical treatment for BPH. Observational studies found that compared to tamsulosin, TZ/DZ/AZ use was associated with slower progression of motor impairment and complications in men with PD (8) and lower incidence of PD in men across four databases in the United States (11, 12), Canada (13), and Denmark (11) in analyses conducted by three independent research groups.

However, a report from Sasane et al. suggested that the decreased PD risk for people using TZ/DZ/AZ *vs*. tamsulosin was owing to tamsulosin increasing the risk of developing PD. (12) Specifically, in a comparison between men taking TZ/DZ/AZ and an untreated propensity-score-matched cohort, they found no difference in PD incidence, whereas a similar comparison between men taking tamsulosin and an untreated cohort found an increased PD incidence in tamsulosin users. (12) These data were interpreted to suggest that tamsulosin increases the risk of PD, and the apparent reduction in risk for men taking TZ/DZ/AZ is the result of an improperly selected comparison group.

However, such a comparison has high risk of confounding by indication. Confounding by indication occurs when the treatment choice is influenced by the outcome in unobserved ways. Prodromal PD can cause urinary dysfunction, (14) which is also a primary indication for treatment with TZ/DZ/AZ or tamsulosin. Any analysis, regardless of form, (15) that fails to consider the unobserved differences will find a spurious relationship between medication use and PD risk, especially in comparisons against men who are untreated or using non-BPH medications. The best, and often only, approach to address this problem is by restricting the sample to only people who meet the indication for treatment, often by making comparisons against those treated with a rival drug, in what is known as the active comparator, new user design. (16)

Consequently, we investigated PD risk with a new comparator group: men using the 5α-reductase inhibitors (5ARI) finasteride or dutasteride. Like α1-adrenergic receptor antagonists, 5ARI are commonly prescribed for BPH. (10) We hypothesized that men taking TZ/DZ/AZ will have reduced hazard of developing PD compared to men using a 5ARI and that the hazard for men taking a 5ARI or tamsulosin will be similar. This comparison creates a new, independent test of both the effect of TZ/DZ/AZ and the suitability of tamsulosin as a comparator group. We tested these hypotheses using a propensity-score-matched cohort of men taking TZ/DZ/AZ, 5ARI, and tamsulosin in a large database of commercial health insurance claims.

## Methods

### Data Source

Health insurance claims for men newly started on terazosin (TZ), doxazosin (DZ), alfuzosin (AZ), tamsulosin, finasteride (5ARI) or dutasteride (5ARI) were obtained from the Truven Analytics Marketscan Commercial Claims and Encounters (CCAE) and Medicare Supplemental and Coordination of Benefits (MDCR) databases for 2001 to 2017. The Marketscan databases contain health insurance claims with diagnoses for all inpatient and outpatient encounters and, for enrollees with prescription drug coverage, claims with the National Drug Code (NDC) number of the dispensed medication. Secondary use of deidentified existing data, such as Truven Marketscan, is not considered human subjects research and is exempt from IRB review.

### Cohort Construction

We identified all prescription events for one of the six medications of interest using the NDC numbers provided for each medication in the 2015 Redbook. For each enrollee, we then identified the first observed dispensing event. We required an enrollee to only take one of the three medication comparison groups and prohibited switching between groups. In other words, an enrollee would only ever take TZ/DZ/AZ, or tamsulosin, or 5ARI.

In order to ensure that the observed dispensing event reflected a newly started therapy, we required at least 365 days of continuous enrollment with prescription drug coverage prior to the index date. We required at least one additional dispensing claim in the year following the first dispensing to ensure we identified actual medication users. As these drugs are rarely used in women, we required that enrollees be male. Both BPH, the most common indication for treatment, and PD are rare diagnoses under the age of 40 and so we required enrollees to be aged at least 40 years at the index date. We required all enrollees to have at least one day of enrollment following the index date and to have been free of PD, see case definition below, on the index date. All enrollees were followed for up to 10 years. Men who left the Truven database or survived for 10 years after medication start date without developing PD were censored at exit or 10 years, whichever occurred first.

### PD Case Definition

We defined a person as having PD if they ever, in any setting, received the diagnosis of PD (ICD-9-CM: 332.0 or ICD-10-CM: G20) or had a prescription drug claim for levodopa. The PD event date was taken as the first appearance of either the diagnosis or prescription dispensing event.

### Propensity Score Matching

Expecting differences between our cohorts, we used propensity score matching to reduce the imbalance. We estimated a propensity score including year of medication start, age, incidence rate of outpatient visits during lookback, mean number of unique diagnoses recorded per outpatient visit during lookback, the incidence rate of diagnoses made in outpatient settings during lookback, incidence rate of inpatient visits during lookback, a history of BPH (ICD-9-CM: 600.xx or ICD-10-CM: N40.x), whether prostate specific antigen (PSA) levels were measured (CPT: 84152, 84153, 84154), if PSA levels were abnormal (ICD-9-CM: 790.93, ICD-10-CM: R97.2, R97.20, R97.21), whether a diagnosis of slow urinary stream was made (ICD-9-CM: 788.62, ICD-10-CM: R39.12), whether a uroflow study was done (ICD-9-CM procedure code: 89.24, ICD-10-CM procedure code: 4A1D75Z, CPT: 51736, 51741), whether a cystometrogram was collected (ICD-9-CM procedure code: 89.22, ICD-10-CM procedure code: 4A0D7BZ, 4A0D8BZ, 4A1D7BZ, 4A1D8BZ, CPT: 51725, 51726), whether a diagnosis of orthostatic hypotension was made (ICD-9-CM: 458.0 or ICD-10-CM: I95.1), a diagnosis of other hypotension (ICD-9-CM: 458.1, 458.2x, 458.8, 458.9 or ICD-10-CM: I95.0, I95.2, I95.3), the 30 Elixhauser comorbidity flags, as revised by AHRQ, with the R package icd. (17, 18)

For year of medication start, we included a series of dummy variables as we expected both changes in medication use patterns and the rate of diagnosis of PD across our sample. Additionally, insurance companies enter and leave the Truven project creating discontinuities in the cohorts. Including dummy variables for year accounts for these problems. We used splines of all the continuous variables to allow for non-linear responses and estimated the propensity score using generalized additive models with a logistic link. Model estimation was performed with the R package mgcv (19, 20) in R 4.0.4. (21)

We used a two-step matching algorithm. First, we required the time from the medication start date to the end of enrollment to be the same (+/-90 days) to ensure balance in time-at-risk between the cases and controls. Second, within the set of possible matches with similar follow-up, we used greedy nearest-neighbor matching based on the estimated log odds (22). To ensure matches were high quality, we imposed a caliper equal to 20% of the pooled standard deviation of the log odds. We matched 1:1 without replacement. In the event of multiple equally good matches, we selected the matching control observation at random. We then had three matched cohorts for the three comparisons of interest: TZ/DZ/AZ versus tamsulosin, TZ/DZ/AZ versus 5ARI, and tamsulosin versus 5ARI.

### Assessing Propensity Score Match Balance

We compared the groups before and after matching on the variables included in the propensity score model. Our primary measure of balance was Cohen’s d, a common measure of effect size, between the groups. We defined the absolute value of Cohen’s d of less than 0.1 as indicating minimal differences between the groups on a covariate. (23, 24)

### Survival Analysis

We visualized the survival curves between the matched groups using a Kaplan Meier survival curve. We tested whether the two curves were different using the non-parametric log-rank test and quantified the difference using semi-parametric Cox proportional hazards regression. For the Cox regression, we used robust standard errors clustered by the pair generated by the propensity score matching. Deviation from the proportional hazards assumption was assessed by comparing the Schoenfeld residuals and time. All survival analysis was done using the R package `survival` (25).

All data and code were checked by the Biostatistics, Epidemiology, and Research Design core as part of the Institute for Clinical and Translational Sciences (ICTS) at the University of Iowa. All code is available at https://github.com/iacobus42/parkinson-disease-5ari.

## Results

After propensity score matching, groups for all three comparisons were well-balanced. All had an absolute value of Cohen’s d<0.10, our pre-specified threshold for balance, with the majority being less than 0.01 (Tables S1-S3).

Our matched cohort size and disease incidence are reported in Table 1. We observed 27.7 cases per 10,000 men per year taking TZ/DZ/AZ compared to 38.5 cases per 10,000 men taking tamsulosin (log rank p-value < 0.0001). Likewise, men taking TZ/DZ/AZ had a lower incidence (24.6 per 10,000 per year) then matched men taking 5ARI (32.4 per 10,000 per year), a difference that was statistically significant (log rank p-value = 0.0006). The difference was smaller and non-significant when comparing men taking tamsulosin, 34.7 cases per 10,000 men per year, to men taking 5ARI, 31.5 cases per 10,000 men per year (p = 0.0512). Kaplan Meier survival curves for all three comparisons are reported in Figure 1.

**Table 1:** Summary of Matched Cohort, Number at Risk, Cases, Incidence Rates, and Log Rank Test. All groups were matched 1:1 based on a propensity score that included age, comorbidities, past medical utilization, and medication start date as well as approximate matching on the duration of post-medication start date enrollment time.

| Comparison | Medication | Number of Enrollees | Person Years of Follow Up Time | Number of Cases of PD | Incidence of PD per 10,000 Person-Years | Log Rank Test p-value |
| --- | --- | --- | --- | --- | --- | --- |
| TZ/DZ/AZ vs. Tamsulosin | TZ/DZ/AZ | 119,973 | 353,634.8 | 973 | 27.5 | $8 \times 10^{-16}$ |
|  | Tamsulosin | 119,973 | 352,698.4 | 1,359 | 38.5 |  |
| TZ/DZ/AZ vs. 5ARI | TZ/DZ/AZ | 64,660 | 193,497.3 | 510 | 26.4 | 0.0006 |
|  | 5ARI | 64,660 | 193,456.0 | 626 | 32.4 |  |
| Tamsulosin vs. 5ARI | Tamsulosin | 78,745 | 238,066.4 | 827 | 34.7 | 0.0512 |
|  | 5ARI | 78,745 | 238,323.6 | 751 | 31.5 |  |

**Figure 1:**
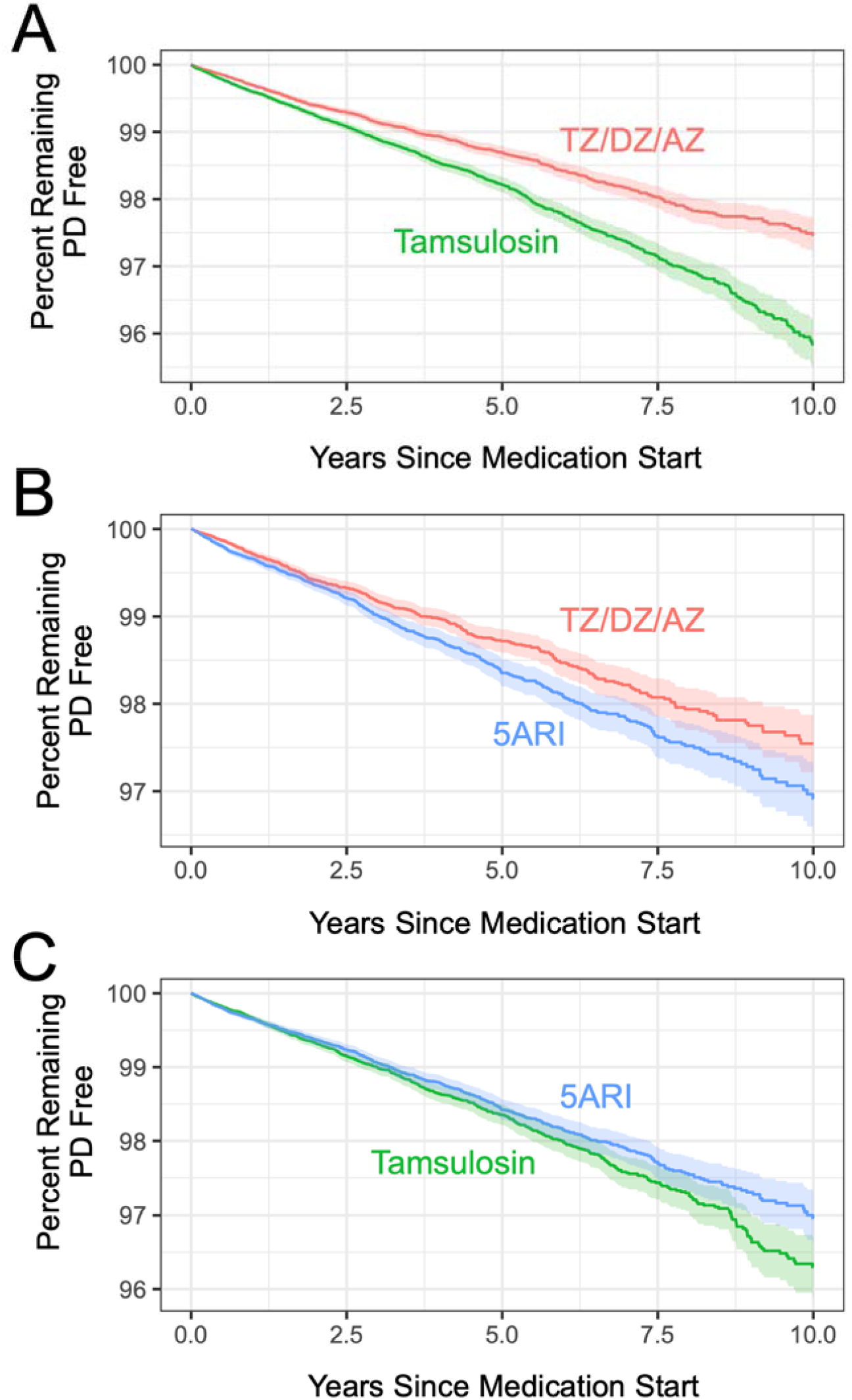
TZ/DZ/AZ decreases risk of PD compared to tamsulosin and 5ARI. Kaplan Meier Survival Curves for matched cohorts A) for TZ/DZ/AZ vs. tamsulosin, B) for TZ/DZ/AZ vs. 5ARI, and C) for tamsulosin vs. 5ARI.

Semi-parametric estimation with Cox proportional hazards regression returned similar results, Table 2. After matching based on propensity scores, we found a significant reduction in the hazard of PD for men taking TZ/DZ/AZ compared to tamsulosin (HR = 0.71, 95% CI: 0.66-0.78). This effect was robust to changing the control group to men taking 5ARI with a 19% reduction in hazard for men who took TZ/DZ/AZ (HR = 0.81, 95% CI: 0.72, 0.92). We failed to find a significant difference in the hazard of PD for men using tamsulosin compared to men taking 5ARI (HR = 1.00, 95% CI: 1.00-1.22). Schoenfeld residuals showed no statistically relevant deviation from the proportional hazards assumption (see supplemental information).

**Table 2:** Results of Cox Proportional Hazards Regression. Propensity-score matched cohorts were matched 1:1 on a propensity score that included age, comorbidities, past medical utilization, and medication start date as well as approximate matching on the duration of post-medication start date enrollment time. Robust standard errors were used and the standard errors were clustered by pair on the matched data set. The second medication in each comparison is the reference group.

| Comparison | Unadjusted |  |  | Propensity Score Matched |  |  |  |
| --- | --- | --- | --- | --- | --- | --- | --- |
|  | Hazard Ratio | 95% CI Lower Bound | 95% CI Upper Bound | Hazard Ratio | 95% CI Lower Bound | 95% CI Upper Bound | p-value |
| TZ/DZ/AZ vs. Tamsulosin | 0.67 | 0.62 | 0.71 | 0.71 | 0.66 | 0.78 | <0.0001 |
| TZ/DZ/AZ vs. 5ARI | 0.88 | 0.80 | 0.97 | 0.81 | 0.72 | 0.92 | 0.0006 |
| Tamsulosin vs. 5ARI | 1.32 | 1.22 | 1.43 | 1.10 | 1.00 | 1.22 | 0.0528 |

## Discussion

This study presents two important findings. First, the use of TZ/DZ/AZ was associated with a decreased hazard of developing PD when compared to a new comparison cohort of men taking 5ARI. This finding is convergent with the initial (11) and replicated result (12, 13) finding lower hazard among men taking TZ/DZ/AZ against tamsulosin. Second, the non-significant difference in hazard, despite an extremely large sample size, between tamsulosin and 5ARI users supports the “null effect” assumption about tamsulosin and the validity of using tamsulosin users as a comparison group.

A key challenge in observational research, and pharmacoepidemiology in particular, is understanding the process that leads to treatment decisions. This is a particular problem when these factors are unobserved, a problem unaddressable by statistical adjustment or propensity-score matching. Comparisons between men taking TZ/DZ/AZ, tamsulosin, or 5ARI are examples of the active comparator design. By conditioning on treatment and, indirectly, indication, we likely reduce some of the residual confounding relative to designs using untreated men as a comparison group. (16)

Beyond simply being men treated for BPH and addressing the potential confounding by indication, men using 5ARIs provide a new comparison group allowing us to directly assess the validity of the initial tamsulosin control. If tamsulosin truly increased the risk of PD and TZ/DZ/AZ truly had no effect, we should see such an effect in this comparison. The absence of such a result suggests the results previously reported based on comparisons against untreated men are spurious.

Our study has some limitations. The relationship between prodromal PD and lower urinary tract symptoms may introduce bias in our comparison against 5ARI. As 5ARIs are used to treat men with less severe symptoms, on average, then men treated with TZ/DZ/AZ or tamsulosin, (10) this is a plausible threat to our results. However, such a bias would tend to make TZ/DZ/AZ or tamsulosin appear worse than they truly are. As this bias would cause us to underestimate the effect, and we still find a significant reduction in hazard, it appears to not be driving our results. While our propensity score model includes many health-related factors and measures specific to the BPH treatment decision, such as diagnosis of abnormal PSA, we do not observe lower urinary tract symptom severity. Other possible factors, such as orthostatic hypotension, appear to be poorly recorded in the diagnostic record. It is plausible that these factors, common to any administrative data analysis, may confound our estimates.

The results of our study, together with four previous retrospective studies from four independent datasets in three countries provide remarkably consistent evidence that use of TZ/DZ/AZ is associated with a decreased risk of developing PD. These results combined with observations that impaired brain energy metabolism may predispose to PD, with the mechanistic discovery that TZ enhances glycolysis and energy metabolism, and with findings that TZ slows or prevents neurodegeneration in multiple toxin-induced and genetic models of PD (8) provide convergent evidence that TZ/DZ/AZ slows neurodegeneration in PD. Nonetheless, a randomized, double-blind, placebo-controlled trial will be required to demonstrate that TZ/DZ/AZ reduces the risk of PD.

## Data Availability

Data is used under license from Truven Analytics/IBM Health.

