## Supplemental for "Use of glycolysis enhancing drugs has less risk of Parkinson’s disease than 5α-reductase inhibitors"

Table S1: Summary Measures of TZ/DZ/AZ Versus Tamsulosin Cohort. Dichotomous variables are reported as precents and continuous variables are reported as the median and inner quartile range. The column d is the Cohen’s d as a measure of difference between the two groups. Rows with absolute values of d greater than 0.1, our threshold for balance, are bolded.

|  | **Before Matching** | | | **After Matching** | | |
| --- | --- | --- | --- | --- | --- | --- |
| **Variable** | **TZ/DZ/AZ** | **Tamsulosin** | **d** | **TZ/DZ/AZ** | **Tamsulosin** | **d** |
| N | 124,905 | 429,741 |  | 119,973 | 119,973 |  |
| Duration of… |  |  |  |  |  |  |
| Lookback in Years | 2.9  (1.7, 5.2) | 3.6  (3.0, 6.3) | -0.24 | 2.94  (1.76, 5.30) | 3.18 (1.88, 5.48) | -0.06 |
| Follow-up in Years | 1.96  (0.93, 4.19) | 1.90  (0.86, 3.78) | 0.09 | 2.05  (0.93, 4.29) | 2.07  (0.92, 4.29) | 0.00 |
| Age | 62  (55, 70) | 62  (56, 71) | -0.05 | 62  (55, 70) | 62  (55, 70) | -0.01 |
| Annual Rate of Inpatient | 0.00  (0.00, 0.12) | 0.00  (0.00, 0.21) | -0.08 | 0.00  (0.00, 0.13) | 0.00  (0.00, 0.14) | -0.01 |
| Annual Rate of Outpatient | 9  (5, 16) | 10  (5, 18) | -0.07 | 9  (5, 16) | 9  (5, 16) | -0.01 |
| Mean Number of Diagnoses | **1.37**  **(1.18, 1.83)** | **1.56**  **(1.27, 2.09)** | **-0.28** | 1.38  (1.19, 1.86) | 1.38  (1.19, 1.87) | -0.01 |
| Annual Rate of Outpatient Diagnoses | **14**  **(7, 25)** | **16**  **(9, 29)** | **-0.13** | 14  (7, 25) | 14  (7, 25) | -0.01 |
| Year of Medication Start | **2009**  **(2006, 2012)** | **2012**  **(2008, 2014)** | **-0.48** | 2009  (2006, 2012) | 2009  (2006, 2012) | -0.01 |
| Percent With BPH Diagnosis | **33.6** | **41.9** | **-0.17** | 34.3 | 35.0 | -0.01 |
| Percent With PSA Measurement | 57.3 | 57.5 | 0.00 | 57.2 | 57.7 | -0.01 |
| Percent With Abnormal PSA | **11.2** | **16.2** | **-0.14** | 11.5 | 11.4 | 0.00 |
| Percent With Slow Urinary Stream | 2.3 | 3.2 | -0.05 | 2.4 | 2.5 | -0.01 |
| Percent with Uroflow Study | 6.0 | 6.5 | -0.02 | 5.2 | 6.2 | 0.00 |
| Percent With Cystometrogram | 0.7 | 0.8 | -0.01 | 0.7 | 0.7 | 0.00 |
| Percent With Orthostatic Hypotension Diagnosis | 0.7 | 1.2 | -0.04 | 0.7 | 0.8 | -0.01 |
| Percent With Other Hypotension Diagnosis | 1.7 | 2.9 | -0.08 | 1.8 | 1.8 | 0.00 |
| Percent With Comorbidity… |  |  |  |  |  |  |
| Alcohol Abuse | 2.1 | 2.6 | -0.03 | 2.2 | 2.2 | 0.00 |
| Anemia | 13.4 | 15.5 | -0.06 | 13.5 | 13.7 | -0.01 |
| Blood Loss | 1.2 | 1.6 | -0.04 | 1.2 | 1.2 | 0.00 |
| Heart Failure | 8.0 | 9.6 | -0.06 | 8.1 | 8.2 | 0.00 |
| Coagulopathy | 2.9 | 4.3 | -0.07 | 2.9 | 3.1 | -0.01 |
| Depression | 7.0 | 9.8 | -0.10 | 7.2 | 7.3 | 0.00 |
| Diabetes (Uncomplicated) | 28.9 | 27.7 | 0.03 | 29.0 | 29.0 | 0.00 |
| Diabetes (Complicated) | 11.3 | 10.6 | 0.02 | 11.3 | 11.4 | 0.00 |
| Drug Abuse | 1.0 | 1.5 | -0.04 | 1.1 | 1.1 | 0.00 |
| Electrolyte Disorders | 11 | 13 | -0.05 | 11.0 | 11.1 | 0.00 |
| HIV | 0.4 | 0.3 | 0.01 | 0.4 | 0.4 | 0.00 |
| Hypertension (Uncomplicated) | **69.4** | **62.0** | **0.15** | 68.9 | 69.3 | -0.01 |
| Hypertension (Complicated) | **16.4** | **12.9** | **0.10** | 16.2 | 16.4 | 0.00 |
| Hypothyroidism | 7.7 | 9.9 | -0.07 | 7.9 | 8.0 | 0.00 |
| Liver Disease | 3.8 | 5.6 | -0.08 | 3.9 | 4.0 | 0.00 |
| Lymphoma | 1.0 | 1.4 | -0.04 | 1.1 | 1.1 | 0.00 |
| Metastatic Cancer | 1.2 | 2.5 | -0.08 | 1.3 | 1.3 | -0.01 |
| Other Neuro. Conditions | 5.8 | 9.2 | -0.12 | 6.0 | 6.1 | -0.01 |
| Obesity | 9.1 | 11 | -0.06 | 9.4 | 9.5 | 0.00 |
| Paralysis | 1.6 | 2.4 | -0.05 | 1.7 | 1.7 | -0.01 |
| Pulmonary Hypertension | 1.9 | 3.0 | -0.07 | 2.0 | 2.1 | -0.01 |
| Psychoses | 5.4 | 7.2 | -0.07 | 5.5 | 5.6 | 0.00 |
| Peptic Ulcer Disease | 0.2 | 0.4 | -0.03 | 0.2 | 0.2 | 0.00 |
| COPD | **19.0** | **23.8** | **-0.12** | 19.4 | 19.5 | 0.00 |
| Peripheral Vascular Disease | 12.0 | 14.6 | -0.08 | 12.2 | 12.4 | 0.00 |
| Renal Failure | 9.5 | 7.1 | 0.09 | 9.4 | 9.5 | 0.00 |
| Rheumatoid Arthritis | 3.7 | 5.0 | -0.06 | 3.8 | 3.8 | 0.00 |
| Solid Tumor | **10.9** | **16.8** | **-0.16** | 11.3 | 11.3 | 0.00 |
| Valvular Disease | 11.1 | 14.1 | -0.09 | 11.3 | 11.5 | -0.01 |
| Weight Loss | 2.7 | 4.5 | -0.09 | 2.7 | 2.9 | -0.01 |

Table S2: Summary Measures of TZ/DZ/AZ Versus 5ARI Cohort. Dichotomous variables are reported as precents and continuous variables are reported as the median and inner quartile range. The column d is the Cohen’s d as a measure of difference between the two groups. Rows with absolute values of d greater than 0.1, our threshold for balance, are bolded.

|  | **Before Matching** | | | **After Matching** | | |
| --- | --- | --- | --- | --- | --- | --- |
| **Variable** | **TZ/DZ/AZ** | **5ARI** | **d** | **TZ/DZ/AZ** | **5ARI** | **d** |
| N | 124,905 | 79,133 |  | 64,660 | 64,660 |  |
| Duration of… |  |  |  |  |  |  |
| Lookback in Years | 2.85  (1.71, 5.18) | 3.15  (1.83, 5.57) | -0.07 | 3.02  (1.80, 5.44) | 3.10  (1.81, 5.46) | -0.01 |
| Follow-up in Years | 1.96  (0.93, 4.19) | 2.23  (0.98, 4.43) | -0.05 | 2.15  (0.94, 4.37) | 2.16  (0.95, 4.37) | 0.00 |
| Age | 62  (55, 70) | 62  (54, 70) | 0.05 | 61  (54, 70) | 61  (54, 70) | -0.01 |
| Annual Rate of Inpatient | **0.00**  **(0.00, 0.12)** | **0.00**  **(0.00, 0.00)** | **0.11** | 0.00  (0.00, 0.00) | 0.00  (0.00, 0.00) | 0.00 |
| Annual Rate of Outpatient | 9  (5, 16) | 9  (5, 16) | 0.05 | 9  (5, 16) | 9  (5, 16) | -0.01 |
| Mean Number of Diagnoses | 1.37  (1.18, 1.83) | 1.40  (1.20, 1.87) | -0.04 | 1.39  (1.19, 1.86) | 1.40  (1.19, 1.88) | -0.02 |
| Annual Rate of Outpatient Diagnoses | 14  (7, 25) | 13  (7, 24) | 0.04 | 13  (7, 24) | 13  (7, 24) | -0.01 |
| Year of Medication Start | **2009**  **(2006, 2012)** | **2010**  **(2008, 2013)** | **-0.22** | 2010  (2007, 2013) | 2010  (2007, 2012) | -0.03 |
| Percent With BPH Diagnosis | **33.6** | **45.0** | **-0.24** | 42.5 | 41.9 | 0.01 |
| Percent With PSA Measurement | 57.3 | 60.7 | -0.07 | 59.0 | 59.7 | -0.01 |
| Percent With Abnormal PSA | **11.2** | **29.7** | **0.49** | 19.9 | 20.6 | 0.01 |
| Percent With Slow Urinary Stream | 2.3 | 2.4 | -0.01 | 2.6 | 2.5 | 0.01 |
| Percent with Uroflow Study | 6.0 | 7.0 | -0.04 | 7.3 | 6.9 | 0.02 |
| Percent With Cystometrogram | 0.7 | 0.7 | 0.00 | 0.8 | 0.7 | 0.01 |
| Percent With Orthostatic Hypotension Diagnosis | 0.7 | 0.9 | -0.03 | 0.8 | 0.9 | 0.00 |
| Percent With Other Hypotension Diagnosis | 1.7 | 1.8 | 0.01 | 1.8 | 1.8 | 0.00 |
| Percent With Comorbidity… |  |  |  |  |  |  |
| Alcohol Abuse | 2.1 | 1.4 | 0.05 | 1.6 | 1.6 | 0.00 |
| Anemia | 13.4 | 11.4 | 0.06 | 11.6 | 11.7 | 0.00 |
| Blood Loss | 1.2 | 1.1 | 0.01 | 1.1 | 1.1 | 0.00 |
| Heart Failure | 8.0 | 6.6 | 0.05 | 6.7 | 6.8 | 0.00 |
| Coagulopathy | 2.9 | 2.9 | 0.00 | 2.9 | 2.9 | 0.00 |
| Depression | 7.0 | 6.8 | 0.01 | 7.1 | 7.0 | 0.01 |
| Diabetes (Uncomplicated) | **28.9** | **19.8** | **0.21** | 21.6 | 21.6 | 0.00 |
| Diabetes (Complicated) | **11.3** | **6.3** | **0.17** | 7.1 | 7.2 | -0.01 |
| Drug Abuse | 1.0 | 0.7 | 0.04 | 0.8 | 0.8 | 0.00 |
| Electrolyte Disorders | **10.8** | **7.3** | **0.12** | 7.9 | 7.9 | 0.00 |
| HIV | 0.4 | 0.7 | 0.04 | 0.6 | 0.6 | 0.00 |
| Hypertension (Uncomplicated) | **69.4** | **52.8** | **0.35** | 56.3 | 56.3 | 0.00 |
| Hypertension (Complicated) | **16.4** | **9.9** | **0.19** | 10.8 | 10.9 | 0.00 |
| Hypothyroidism | 7.7 | 8.8 | -0.04 | 8.4 | 8.6 | -0.01 |
| Liver Disease | 3.8 | 3.1 | 0.04 | 3.3 | 3.3 | 0.00 |
| Lymphoma | 1.0 | 1.0 | 0.00 | 1.1 | 1.0 | 0.00 |
| Metastatic Cancer | 1.2 | 1.4 | -0.01 | 1.4 | 1.3 | 0.01 |
| Other Neuro. Conditions | 5.8 | 5.6 | 0.01 | 1.4 | 1.3 | 0.01 |
| Obesity | 9.1 | 5.6 | 0.13 | 6.3 | 6.4 | -0.01 |
| Paralysis | 1.6 | 1.2 | 0.03 | 1.3 | 1.3 | 0.00 |
| Pulmonary Hypertension | 1.9 | 1.9 | 0.00 | 1.9 | 1.9 | 0.00 |
| Psychoses | 5.4 | 5.2 | 0.01 | 5.4 | 5.3 | 0.00 |
| Peptic Ulcer Disease | 0.2 | 0.2 | 0.00 | 0.2 | 0.2 | 0.00 |
| COPD | 19.0 | 17.7 | 0.03 | 18.1 | 18.0 | 0.00 |
| Peripheral Vascular Disease | 12.0 | 10.4 | 0.05 | 10.7 | 10.8 | 0.00 |
| Renal Failure | **9.5** | **4.3** | **0.20** | 4.9 | 5.0 | -0.01 |
| Rheumatoid Arthritis | 3.7 | 3.5 | 0.01 | 3.6 | 3.6 | 0.00 |
| Solid Tumor | 10.9 | 13.1 | -0.07 | 12.4 | 12.3 | 0.00 |
| Valvular Disease | 11.1 | 11.5 | -0.01 | 11.4 | 11.2 | 0.00 |
| Weight Loss | 2.7 | 2.8 | -0.01 | 2.8 | 2.8 | 0.00 |

Table S3: Summary Measures of Tamsulosin Versus 5ARI Cohort. Dichotomous variables are reported as precents and continuous variables are reported as the median and inner quartile range. The column d is the Cohen’s d as a measure of difference between the two groups. Rows with absolute values of d greater than 0.1, our threshold for balance, are bolded.

|  | **Before Matching** | | | **After Matching** | | |
| --- | --- | --- | --- | --- | --- | --- |
| **Variable** | **Tamsulosin** | **5ARI** | **d** | **Tamsulosin** | **5ARI** | **d** |
| N | 429,741 | 79,133 |  | 78,745 | 78,745 |  |
| Duration of… |  |  |  |  |  |  |
| Lookback in Years | **3.6**  **(2.0, 6.3)** | **3.1**  **(1.8, 5.6)** | **0.18** | 3.31  (1.92, 5.78) | 3.15  (1.84, 5.57) | 0.06 |
| Follow-up in Years | **1.90**  **(0.86, 3.78)** | **2.23**  **(0.98, 4.43)** | **-0.14** | 2.23  (0.98, 4.41) | 2.22  (0.98, 4.41) | 0.00 |
| Age | **62**  **(56, 71)** | **62**  **(54, 70)** | **0.10** | 62  (54, 71) | 62  (54, 71) | 0.00 |
| Annual Rate of Inpatient | **0.00**  **(0.00, 0.21)** | **0.00**  **(0.00, 0.00)** | **0.18** | 0.00  (0.00, 0.00) | 0.00  (0.00, 0.00) | 0.02 |
| Annual Rate of Outpatient | **10**  **(5, 18)** | **9**  **(5, 16)** | **0.11** | 9  (5, 16) | 9  (5, 16) | 0.01 |
| Mean Number of Diagnoses | **1.56**  **(1.27, 2.09)** | **1.40**  **(1.20, 1.87)** | **0.24** | 1.41  (1.20, 1.88) | 1.41  (1.20, 1.87) | 0.01 |
| Annual Rate of Outpatient Diagnoses | **16**  **(9, 29)** | **13**  **(7, 24)** | **0.16** | 13  (7, 24) | 13  (7, 24) | 0.02 |
| Year of Medication Start | **2012**  **(2008, 2014)** | **2010**  **(2008, 2013)** | **0.27** | 2010  (2008, 2013) | 2010  (2008, 2013) | 0.01 |
| Percent With BPH Diagnosis | 41.9 | 45.0 | -0.06 | 45.0 | 44.9 | 0.00 |
| Percent With PSA Measurement | 57.5 | 60.7 | -0.07 | 60.7 | 60.6 | 0.00 |
| Percent With Abnormal PSA | **16.2** | **29.7** | **-0.35** | 29.7 | 29.5 | 0.00 |
| Percent With Slow Urinary Stream | 3.2 | 2.4 | 0.05 | 2.5 | 2.5 | 0.00 |
| Percent with Uroflow Study | 6.5 | 7.0 | -0.02 | 7.3 | 7.0 | 0.01 |
| Percent With Cystometrogram | 0.8 | 0.7 | 0.01 | 0.7 | 0.7 | 0.01 |
| Percent With Orthostatic Hypotension Diagnosis | 1.2 | 0.9 | 0.02 | 1.0 | 0.9 | 0.00 |
| Percent With Other Hypotension Diagnosis | 2.9 | 1.8 | 0.07 | 1.9 | 1.8 | 0.00 |
| Percent With Comorbidity… |  |  |  |  |  |  |
| Alcohol Abuse | 2.6 | 1.4 | 0.08 | 1.5 | 1.4 | 0.01 |
| Anemia | 15.5 | 11.4 | 0.12 | 11.6 | 11.5 | 0.01 |
| Blood Loss | 1.6 | 1.1 | 0.04 | 1.1 | 1.1 | 0.00 |
| Heart Failure | 9.6 | 6.6 | 0.11 | 6.8 | 6.6 | 0.01 |
| Coagulopathy | 4.3 | 2.9 | 0.07 | 3.0 | 2.9 | 0.01 |
| Depression | 9.8 | 6.8 | 0.11 | 6.9 | 6.8 | 0.00 |
| Diabetes (Uncomplicated) | **27.7** | **19.8** | **0.18** | 20.4 | 19.9 | 0.01 |
| Diabetes (Complicated) | **10.6** | **6.3** | **0.14** | 6.7 | 6.4 | 0.02 |
| Drug Abuse | 1.5 | 0.7 | 0.07 | 0.8 | 0.7 | 0.01 |
| Electrolyte Disorders | **12.6** | **7.3** | **0.16** | 0.8 | 0.7 | 0.01 |
| HIV | 0.3 | 0.7 | -0.06 | 0.7 | 0.7 | 0.00 |
| Hypertension (Uncomplicated) | **62.0** | **52.8** | **0.19** | 53.1 | 52.9 | 0.00 |
| Hypertension (Complicated) | 12.9 | 9.9 | 0.09 | 10.2 | 9.9 | 0.01 |
| Hypothyroidism | 9.9 | 8.8 | 0.04 | 9.0 | 8.8 | 0.01 |
| Liver Disease | **5.6** | **3.1** | **0.11** | 3.3 | 3.1 | 0.01 |
| Lymphoma | 1.4 | 1.0 | 0.03 | 1.1 | 1.1 | 0.01 |
| Metastatic Cancer | 2.5 | 1.4 | 0.07 | 1.5 | 1.4 | 0.01 |
| Other Neuro. Conditions | **9.2** | **5.6** | **0.13** | 5.8 | 5.6 | 0.01 |
| Obesity | **11.0** | **5.6** | **0.18** | 6.0 | 5.7 | 0.01 |
| Paralysis | 2.4 | 1.2 | 0.08 | 1.3 | 1.2 | 0.01 |
| Pulmonary Hypertension | 3.0 | 1.9 | 0.07 | 2.1 | 1.9 | 0.01 |
| Psychoses | 7.2 | 5.2 | 0.08 | 5.3 | 5.2 | 0.00 |
| Peptic Ulcer Disease | 0.4 | 0.2 | 0.03 | 0.3 | 0.2 | 0.01 |
| COPD | **23.8** | **17.7** | **0.15** | 18.0 | 17.7 | 0.01 |
| Peripheral Vascular Disease | **14.6** | **10.4** | **0.12** | 10.8 | 10.5 | 0.01 |
| Renal Failure | **7.1** | **4.3** | **0.11** | 4.5 | 4.4 | 0.01 |
| Rheumatoid Arthritis | 5.0 | 3.5 | 0.07 | 3.7 | 3.5 | 0.01 |
| Solid Tumor | 16.8 | 13.1 | 0.10 | 14.0 | 13.2 | 0.02 |
| Valvular Disease | 14.1 | 11.5 | 0.08 | 11.8 | 11.5 | 0.01 |
| Weight Loss | 4.5 | 2.8 | 0.08 | 2.9 | 2.8 | 0.01 |

Figure S1: Schoenfeld Residuals for the Matched Cohort. The dashed line and shaded region are a LOESS smooth of the residuals and 95% CI. Schoenfeld residuals did not show concerning deviations from the proportional hazards assumption. We found near zero correlations between time for the Schoenfeld residual for the comparison (TZ/DZ/AZ versus tamsulosin: r = -0.059, p = 0.004; TZ/DZ/AZ versus 5ARI: r = 0.005, p = 0.870; tamsulosin versus 5ARI: r = 0.04, p = 0.080) (Fig. S1). While the correlation was statistically significant for the comparison of TZ/DZ/AZ and tamsulosin, given the very large sample size the test of the residuals is certainly overpowered to detect non-statistically relevant deviations from the proportional hazards assumption.
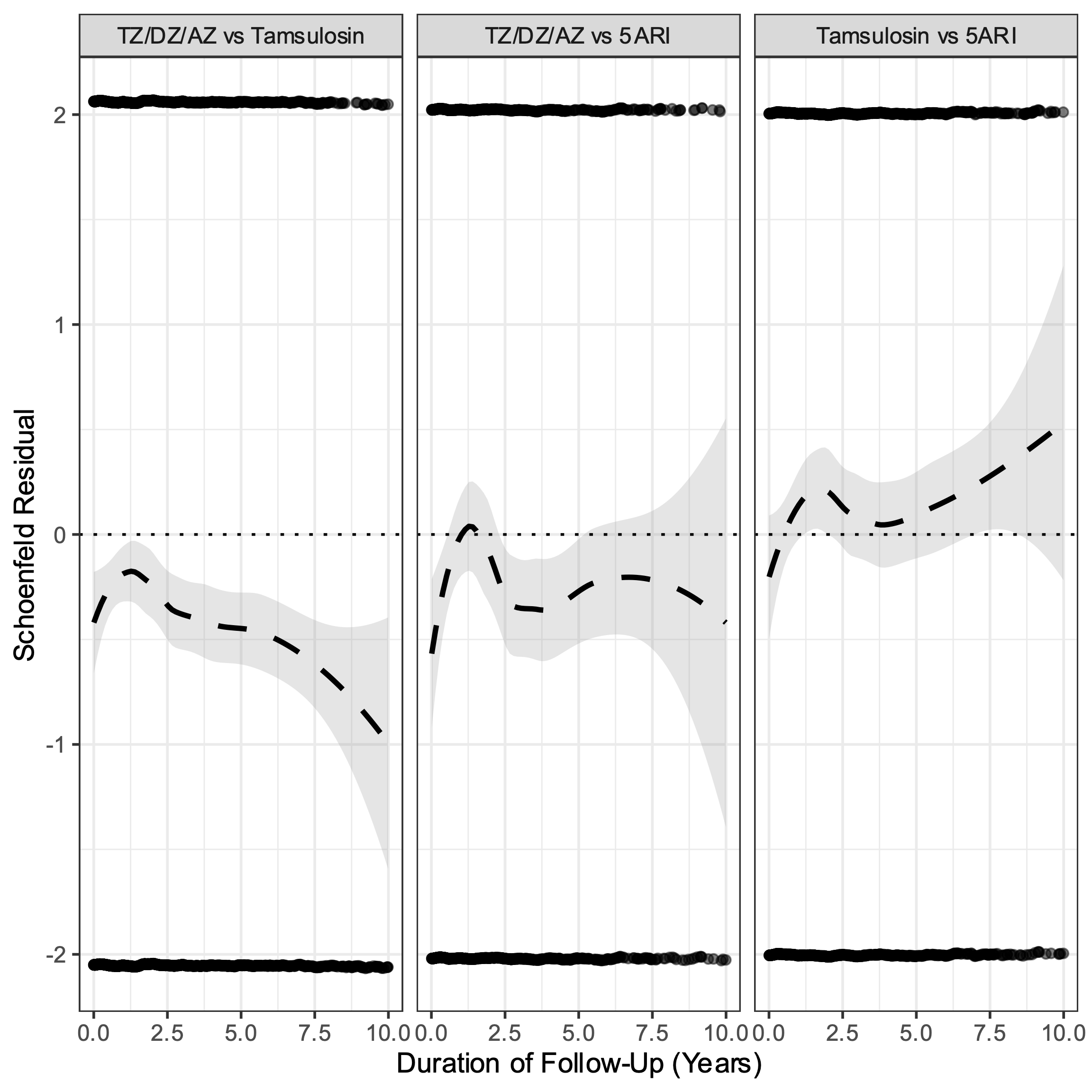
